# Incidence of bacterial sexually transmitted infections and associated factors in men who have sex with men: results from the ANRS 12 400/DepIST-H cohort

**DOI:** 10.64898/2026.07.28.26359109

**Authors:** Oumarou I. Wone Adama, Béatrice Berçot, Arnold Junior Sadio, Fifonsi Adjidossi Gbeasor-Komlanvi, Martin Kouame Tchankoni, Yao Rodion Konu, Mounerou Salou, Ephrem Mensah, Anoumou Claver Dagnra, Jade Ghosn, Valentine Marie Ferré, Diane Descamps, Charlotte Charpentier, Didier Koumavi Ekouevi, Cécile Bébéar

## Abstract

**Background:** The objective was through an analysis of longitudinal data from the DepIST-H cohort to estimate the incidence, describe the dynamics and factors associated with CT/NG infection in MSM in Togo.

**Methods:** A total of 200 MSM in Lomé, Togo, were included in the ANRS I MIE 12400/DepIST-H cohort, half living with HIV. After inclusion in the study, participants received follow-up clinical visits at 6 and 18 months, and laboratory tests were performed at baseline (M0) and at 12 and 24 months (M12 and M24). PCR tests for CT and NG were performed from pharyngeal, anal and urine samples collected using the Xpert® CT/GC kit (Sunnyvale, CA, USA). To identify the factors associated with anal bacterial STIs, offset term Poisson regression models were performed.

**Results:** Among 200 MSM (median age 24 years), CT and NG incidence rates were high (≈22 cases per 100 person-years), particularly at the anal site. Multiple partners significantly increased anal NG incidence (aIRR 2.35). Anal NG incidence was significantly lower among participants with income-generating activity (aIRR 0.57). Trends suggested higher anal CT and NG incidence in the presence of pharyngeal infection, but associations were not statistically significant

**Conclusion:** This study confirms that MSM remain highly exposed to STIs in Togo and highlights the need to strengthen prevention efforts within this population. Postexposure doxycycline prophylaxis and multisite pooled-sample screening represent promising avenues to strengthen prevention efforts within these populations.

## Introduction

Sexually transmitted infections (STIs) are a global public health problem, with more than one million new infections estimated every day among people aged 15 to 49 [1]. According to World Health Organization (WHO) estimates, the number of new cases of curable STIs (chlamydia, gonorrhea, syphilis and trichomoniasis) in 2020 was estimated at about 374 million; *Neisseria gonorrhoeae* (NG) and *Chlamydia trachomatis* (CT) being the most prevalent, with 78 million and 130 million cases respectively [1]. These STIs increase the risk of HIV transmission, making their integration into combination prevention strategies essential [2]. STIs are more common among men who have sex with men (MSM) because of the risky behaviors and stigma they are exposed to [3]. A systematic review and meta-analysis published in 2022 estimated the prevalence of bacterial STIs among MSM living in Latin America to be around 23.9% and an incidence rate of 72.2 cases per 100 person-years [4]. In the United States, the same trends have been observed and recent models suggest that nearly 10% of new HIV infections in this population are attributable to gonorrhea and chlamydia [5].

In sub-Saharan Africa, data on STIs are limited. However, a few studies have been carried out among MSM in West Africa and have reported high prevalences [6, 7]. In the majority of these countries, homosexuality is criminalized, which is a barrier to access to HIV/STI testing and treatment services and limits research among this population [8]. In Togo, the analysis of data at the inclusion of the DepIST-H project, reported a prevalence of 32.5% for NG and 32.0% for CT and a different distribution according to anatomical sites (anus, throat and urinary tract) [7]. The prevalences of NG (22% vs. 19% throat and 3% urine) and CT (26% vs. 8% throat and 5% urine) at the anal level were the highest [7]. This predominance of carrying at the anal level has been confirmed in MSM in several studies, particularly in Kenya, Spain and Canada [9]. In addition, undetected pharyngeal and rectal infections in sexually active MSM may be reservoirs of transmissibility in countries where the syndromic approach is used in the management of STIs. It is therefore crucial to monitor the dynamics of these infections at the anal, pharyngeal and urinary sites to better understand their role in STI transmission and to effectively guide prevention and management strategies. The objective was through an analysis of longitudinal data from the DepIST-H cohort to estimate the incidence, describe the dynamics and factors associated with CT/NG infection in MSM in Togo.

## Methods

### Study design and period

This is an analysis of longitudinal data from the ANRS 12400/DepIST-H cohort. This is a cohort of 200 MSM (100 HIV+ and 100 HIV-) followed over two years in Togo. It was implemented between July 2021 and January 2024 and aimed to estimate the prevalence and incidence of HPV infections and anal lesions [7].

### Study population

The study population consisted of individuals who met the following criteria: (i) self-identify as MSM; (ii) be 18 years of age or older; (iii) have provided informed consent to participate in the study. The participants were recruited at the Centre Espoir Vie Togo (EVT), a community-based facility specializing in HIV care and support for key populations. Recruitment was carried out by the peer educators and health mediators of the EVT.

### Cohort follow-up

After inclusion in the study, participants received follow-up clinical visits at 6 and 18 months, and laboratory tests were performed at baseline (M0) and at 12 and 24 months (M12 and M24). The data collected focused mainly on: (i) risky sexual behaviors; (ii) the results of the clinical examination on STIs; and (iii) biological data, including HIV and bacterial STI testing (CT and NG).

### Data collected during follow-up

#### Behavioral and clinical data

Information on sexual practices, number of casual partners, knowledge of the partners’ HIV status, history of STIs and symptoms suggestive of STIs in the past six months was collected using a digitized questionnaire. For participants who had difficulty expressing themselves in French, the questionnaire was administered in English or local languages to facilitate exchanges. The clinical examination and anal swabs were performed by a physician trained in the diagnosis and management of anal lesions.

#### Screening for Chlamydia trachomatis and Neisseria gonorrhoeae

PCR tests for CT and NG were performed from pharyngeal, anal and urine samples collected using the Xpert® CT/GC kit (Sunnyvale, CA, USA). Positive samples were sent to the National Reference Center for LGV typing (C. trachomatis) and resistance testing (N. gonorrhoeae).

#### HIV testing

Blood samples were taken for HIV testing using the *BIOLINE HIV® (Abbott, Santa Clara, CA, USA), Bioline™ HCV (Abbott) and Alere Determine™ HBsAg (Abbott)* tests. Any positive HIV test was confirmed by a second rapid test, the *First Response® HIV 1–2. O Card Test (Premier Medical, Maharashtra, India),* in accordance with national guidelines. In case of discordant results, the samples were analyzed using the *INNO-LIA® HIV I/II Score immunoimprint (Fujirebio, Gothenburg, Sweden)*.

### Operational definitions

A prevalent infection was defined by a positive test for Chlamydia trachomatis (CT) or Neisseria gonorrhoeae (NG) at the inclusion visit (M0). Regarding incident infections, two approaches were chosen: i) Occurrence of a first positive result at follow-up in participants who were negative at baseline; ii) Any positive results observed after a previous negative visit. This second approach makes it possible to include participants who were positive at inclusion (M0), who then became negative (e.g. at M12), then tested positive again (e.g. at M24).

### Statistical analysis

Descriptive statistics were produced and the results were presented with tables of numbers and proportions for the qualitative variables. Quantitative variables were presented as medians with their interquartile range (IQR). Median comparisons were made using the Wilcoxon– Mann-Whitney test, while proportion comparisons used the Pearson chi-square test. Incidence rates were expressed as the number of events per 100 person-years. To identify the factors associated with anal bacterial STIs, offset term Poisson regression models were performed, using the two definitions of incidence. All statistical analyses were carried out with the R software, version 4.5.1 (R Foundation for Statistical Computing, Vienna, Austria).

### Ethical considerations

The research protocol was approved by the Bioethics Committee for Health Research of the Ministry of Health of Togo (CBRS) under number 039/2019/CBRS. All participants provided written informed consent prior to inclusion in the study. In addition, the study was registered on the International Clinical Trials Registry (ClinicalTrials.gov NLM) under the number NCT04910438.

## Results

### Participant flow diagram from inclusion to last follow-up visit

A total of 200 MSM were included in the study. During the first year of follow-up, two deaths were reported and 16 participants were initially classified as lost to follow-up, ten of whom were difficult to reach and six of whom were travelling outside the city of Lomé. Six of them were eventually found, bringing to 188 the number of participants seen at the M12 visit. During the second year of follow-up, an additional 30 participants were reported to be lost to follow-up and could not be present at the last follow-up visit (Figure 1).

**Figure 1.**
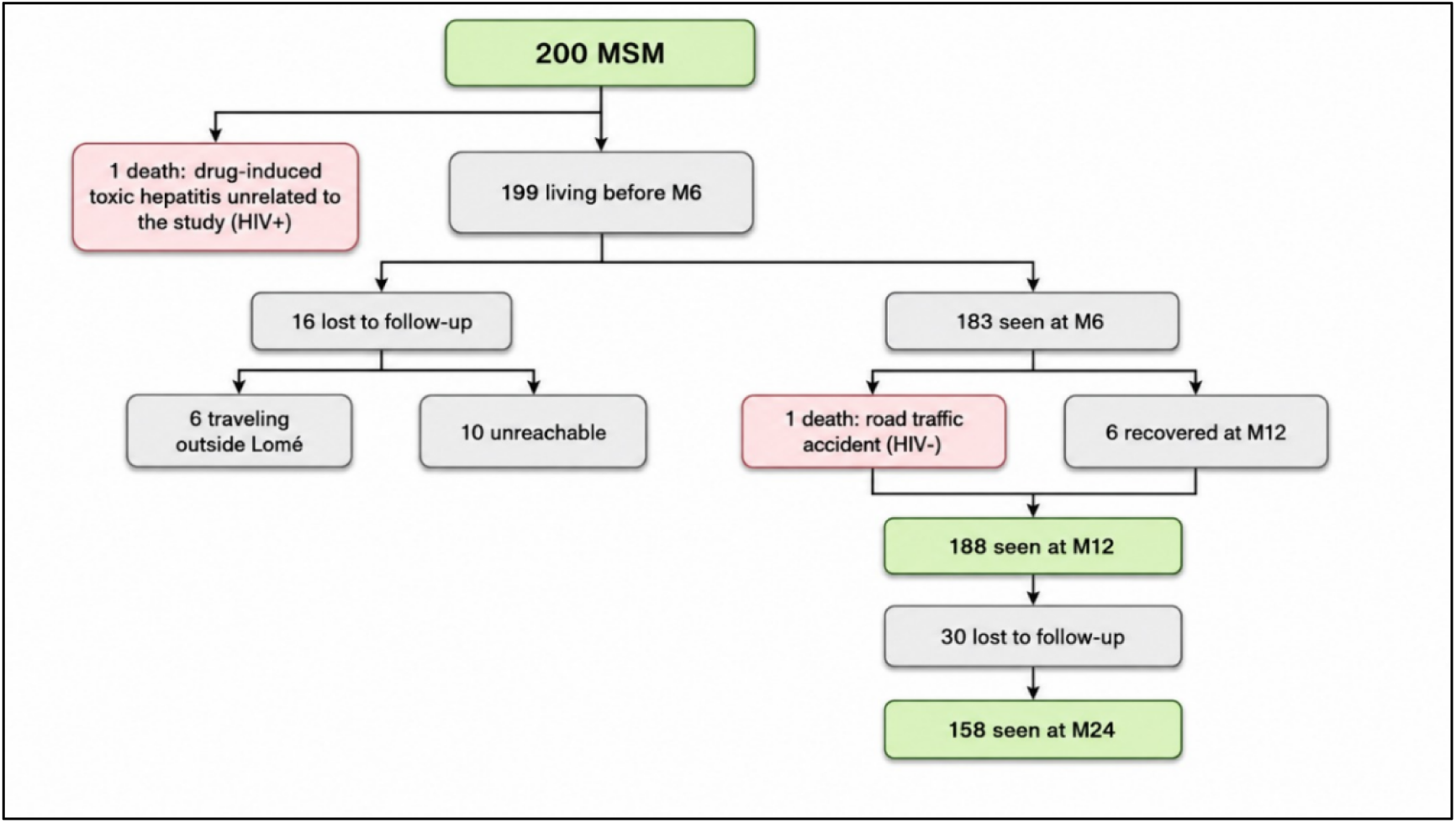
Flow diagram cohort ANRS I MIE 12 400/DepIST-H

### Characteristics of study participants

The median age of the participants was 24 years (IQR: 21–29) and the majority (90.5%) of the included MSM lived alone. Participants living with HIV had a significantly older age than their HIV-negative counterparts (27 years vs. 23 years). Socioeconomically, 38% had no source of income and 40% reported transactional sex. Regarding the number of sexual partners, 75% of participants reported having at least two partners, and 20% reported a practice of chemsex. Table 1 presents the characteristics of the participants’ inclusion.

**Table 1.** Characteristics of MSM at baseline by HIV status, ANRS-12400/DepIST-H cohort.

|  | HIV status |  |  | p-value |
| --- | --- | --- | --- | --- |
|  | Negative<br>N = 100 | Positive<br>N = 100 | Total<br>N = 200 |  |
| **Ages (years)** |  |  |  | <0.001<sup>1</sup> |
| Min-Max | 18-38 | 19-49 | 18-49 |  |
| Median (Q1-Q3) | 22 (20-25) | 27 (23-30) | 24 (21-29) |  |
| **Age groups (years), n (%)** |  |  |  | <0.001<sup>2</sup> |
| <25 | 70 (70.0) | 35 (35.0) | 105 (52.5) |  |
| 25+ | 30 (30.0) | 65 (65.0) | 95 (47.5) |  |
| **Level of education, n (%)** |  |  |  | 0.179<sup>3</sup> |
| Primary | 3 (3.0) | 5 (5.0) | 8 (4.0) |  |
| Secondary (Middle School/High School) | 45 (45.0) | 56 (56.0) | 101 (50.5) |  |
| Higher Education (University) | 52 (52.0) | 39 (39.0) | 91 (45.5) |  |
| **Marital status, n (%)** |  |  |  | 0.138<sup>2</sup> |
| As a couple | 12 (12.0) | 6 (6.0) | 18 (9.0) |  |
| Lives alone | 88 (88.0) | 94 (94.0) | 182 (91.0) |  |
| **Income-generating activity, n (%)** |  |  |  | 0.020<sup>2</sup> |
| Not | 46 (46.0) | 30 (30.0) | 76 (38.0) |  |
| Yes | 54 (54.0) | 70 (70.0) | 124 (62.0) |  |
| **Condom use, n (%)** |  |  |  | 0.777<sup>2</sup> |
| Difficult | 45 (45.0) | 47 (47.0) | 92 (46.0) |  |
| Easy | 55 (55.0) | 53 (53.0) | 108 (54.0) |  |
| **Transactional sex, n (%)** |  |  |  | 0.337<sup>2</sup> |
| Not | 62 (63.3) | 56 (56.6) | 118 (59.9) |  |
| Yes | 36 (36.7) | 43 (43.4) | 79 (40.1) |  |
| **Number of sexual partners, n (%)** |  |  |  | 0.869<sup>2</sup> |
| 1 | 24 (24.0) | 25 (25.0) | 49 (24.5) |  |
| 2+ | 76 (76.0) | 75 (75.0) | 151 (75.5) |  |
| **Anal sex type, n (%)** |  |  |  | 0.027<sup>2</sup> |
| Active | 20 (20.0) | 7 (7.0) | 27 (13.5) |  |
| Passive | 49 (49.0) | 57 (57.0) | 106 (53.0) |  |
| Versatile | 31 (31.0) | 36 (36.0) | 67 (33.5) |  |
| **Practice of chemsex, n (%)** |  |  |  | 0.289<sup>2</sup> |
| Not | 77 (77.0) | 83 (83.0) | 160 (80.0) |  |
| Yes | 23 (23.0) | 17 (17.0) | 40 (20.0) |  |

<sup>1</sup>Wilcoxon rank sum test<sup>2</sup>Pearson's Chi-squared test<sup>3</sup>Fisher's exact test

### Site-specific incidence rates of bacterial STIs in MSM

The incidence rate of CT infection was 21.8 cases per 100 person-years, and the incidence rate of NG was 22.0 cases per 100 person-years. The incidence rate was higher at the anal level with 17.2 cases per 100 person-years for CT and 17.9 cases per 100 person-years for NG (Table 2).

**Table 2.**
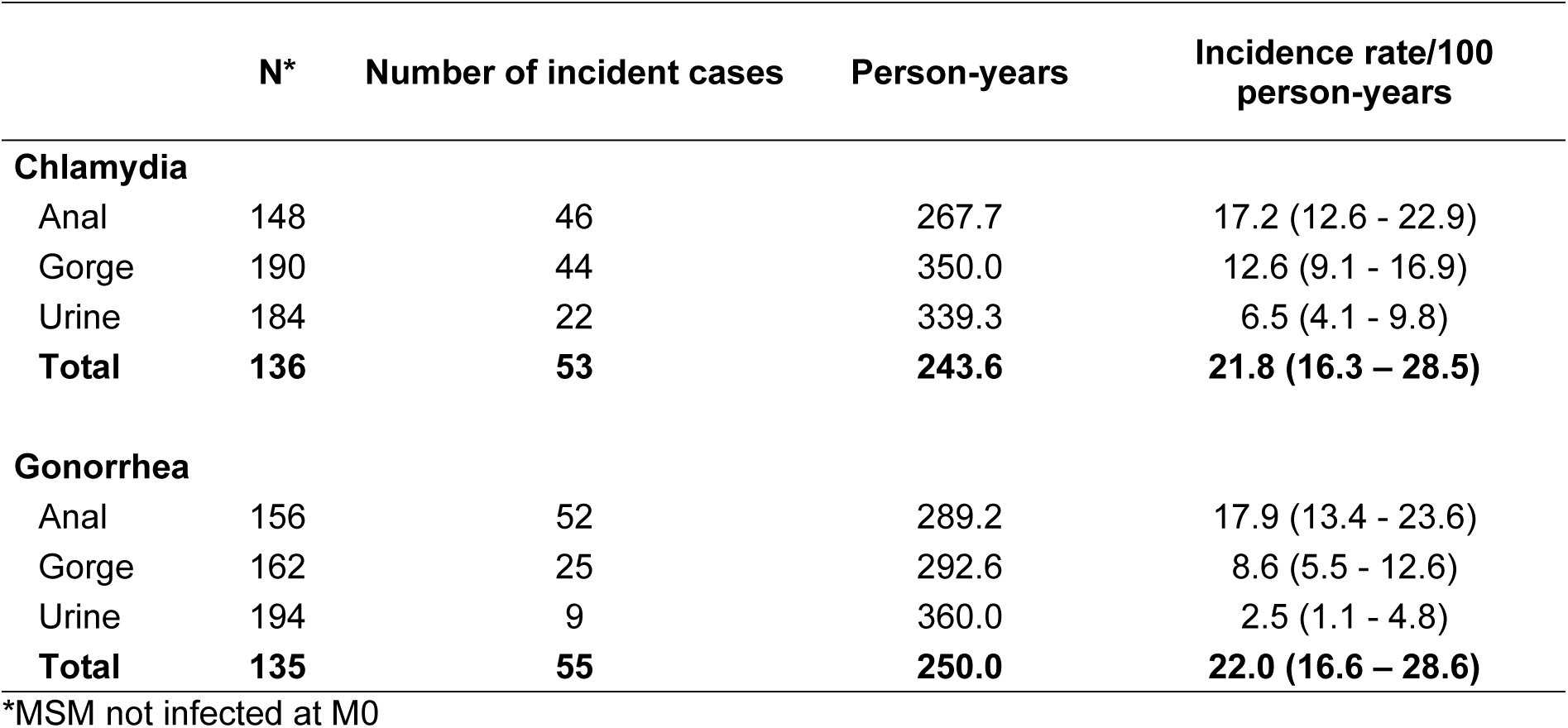
Incidence rate of CT, NG without prevalent infection according to anatomical sites, ANRS Cohort, DepIST H 12400, Togo, 2024.

### Evolution of CT and NG infections at the anal level during the two years of follow-up

Figure 2 describes the evolution of CT infection at the anal level during the two years of follow-up. At baseline, 52 cases of CT were detected at the anal level, then 50 at M12, and 34 at the last visit.

**Figure 2.**
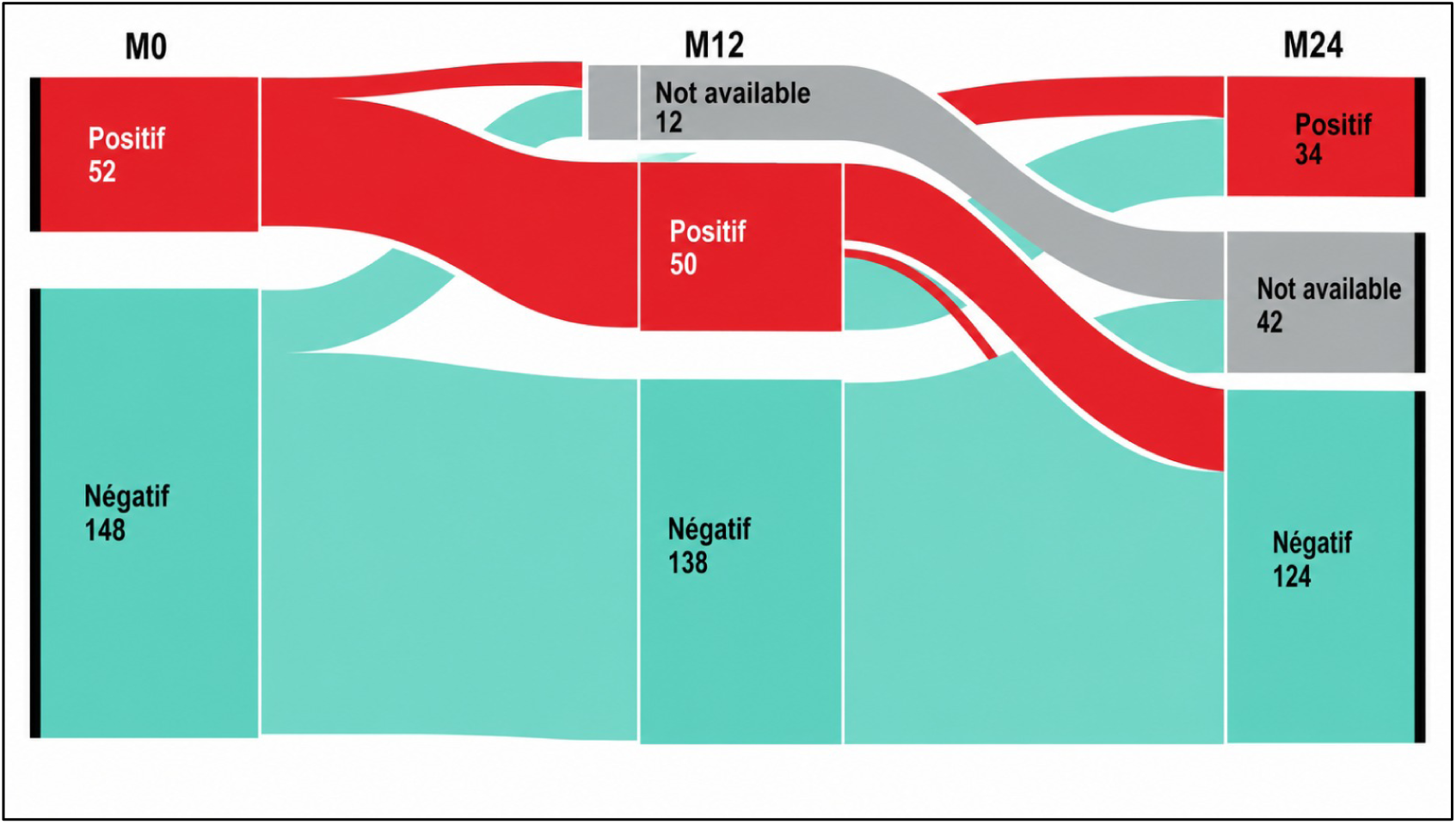
Evolution of CT infections at the anal level during the 2 years of follow-up

The same trends were observed for NG at the anal level. At M0, 44 cases of NG were detected at the anal level, then 41 at M12, and 30 at the last visit (Figure 3).

**Figure 3.**
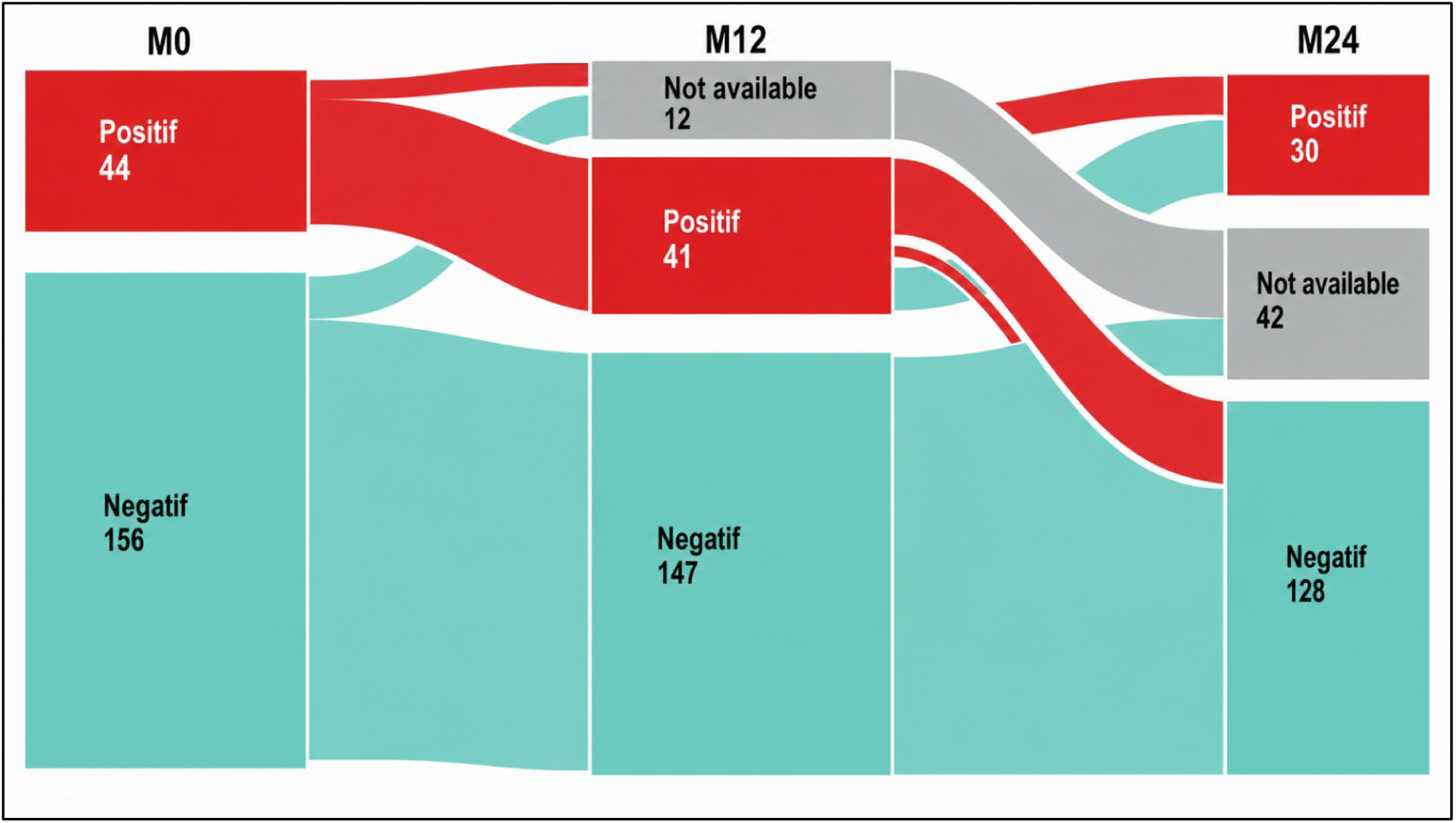
Evolution of NG infections at the anal level during the 2 years of follow-up

### Factors associated with anal CT

The results of Model 1 suggest a trend towards an increased incidence of anal CT in multiple partners (aIRR: 1.34; 95% CI: 0.66 – 2.96), but not statistically significant. In Model 2, a similar trend was noted: the incidence rate appears to be higher overall in the presence of pharyngeal infection (aIRR: 2.96; 95% CI: 0.73 – 8.78) and transactional reports (aIRR: 1.95; 95% CI: 0.89 – 4.30), but wide confidence intervals including 1 do not support a significant association.

**Table 3.** Factors associated with CT at the anal level, ANRS Cohort, DepIST H 12400, Togo, 2024.

|  | Model 1 |  |  | Model 2 |  |  |
| --- | --- | --- | --- | --- | --- | --- |
|  | Multivariate model |  |  | Multivariate model |  |  |
|  | IRRa | IC95% | p | IRRa | IC95% | p |
| **Age groups (years)** |  |  | 0.605 |  |  | 0.484 |
| <25 | 1 | — |  | 1 | — |  |
| 25 + | 1.22 | 0.57 – 2.63 |  | 1.37 | 0.56 – 3.36 |  |
| **HIV status** |  |  | 0.507 |  |  | 0.244 |
| Negative | 1 | — |  | 1 | — |  |
| Positive | 1.26 | 0.65 – 2.51 |  | 0.63 | 0.28 – 1.37 |  |
| **Income-generating activity** |  |  | 0.272 |  |  | 0.183 |
| Not | 1 | — |  | 1 | — |  |
| Yes | 0.66 | 0.33 – 1.39 |  | 0.56 | 0.24 – 1.31 |  |
| **Type of anal intercourse** |  |  | 0.581 |  |  | 0.553 |
| Active | 1 | — |  | 1 | — |  |
| Passive or versatile | 1.31 | 0.53 – 3.97 |  | 0.71 | 0.26 – 2.40 |  |
| **Transactional Sex** |  |  | 0.908 |  |  | 0.094 |
| Not | 1 | — |  | 1 | — |  |
| Yes | 1.04 | 0.53 – 2.01 |  | 1.95 | 0.89 – 4.30 |  |
| **Number of sexual partners** |  |  | 0.433 |  |  | 0.346 |
| 1 | 1 | — |  | 1 | — |  |
| 2+ | 1.34 | 0.66 – 2.96 |  | 1.45 | 0.67 – 3.27 |  |
| **Condom use** |  |  | 0.571 |  |  | 0.427 |
| Difficult | 1 | — |  | 1 | — |  |
| Easy | 0.84 | 0.46 – 1.55 |  | 1.35 | 0.64 – 2.93 |  |
| **Infection CT gorge** |  |  | 0.612 |  |  | 0.115 |
| Not | 1 | — |  | 1 | — |  |
| Yes | 1.92 | 0.15 – 3.16 |  | 2.96 | 0.73 – 8.78 |  |
| **Chemsex Practice** |  |  | 0.645 |  |  | 0.478 |
| Not | 1 | — |  | 1 | — |  |
| Yes | 1.18 | 0.56 – 2.34 |  | 0.71 | 0.24 – 1.76 |  |

95% CI = confidence interval,
IRR = incidence rate ratio
aIRR = Adjusted Incidence Rate Ratio
**Model 1:** incidence defined as the occurrence of a first positive CT or NG result at a follow-up visit after baseline**Model 2:** incidence defined as any positive CT or NG result observed at a visit, after a negative result at the previous visit

### Factors associated with anal NG

The results of Model 1 suggest a significant increase in the incidence of anal NG in the case of multiple partners (aIRR: 2.35; [1,04–6,30]). In Model 2, the incidence rate of NG at the anal level was significantly halved in participants with income-generating activity (aIRR: 0.57; p=0.041). Overall, the results of the two models suggest a trend towards an increase in the incidence of anal NG in the event of pharyngeal infection but are not significant.

**Table 4.**
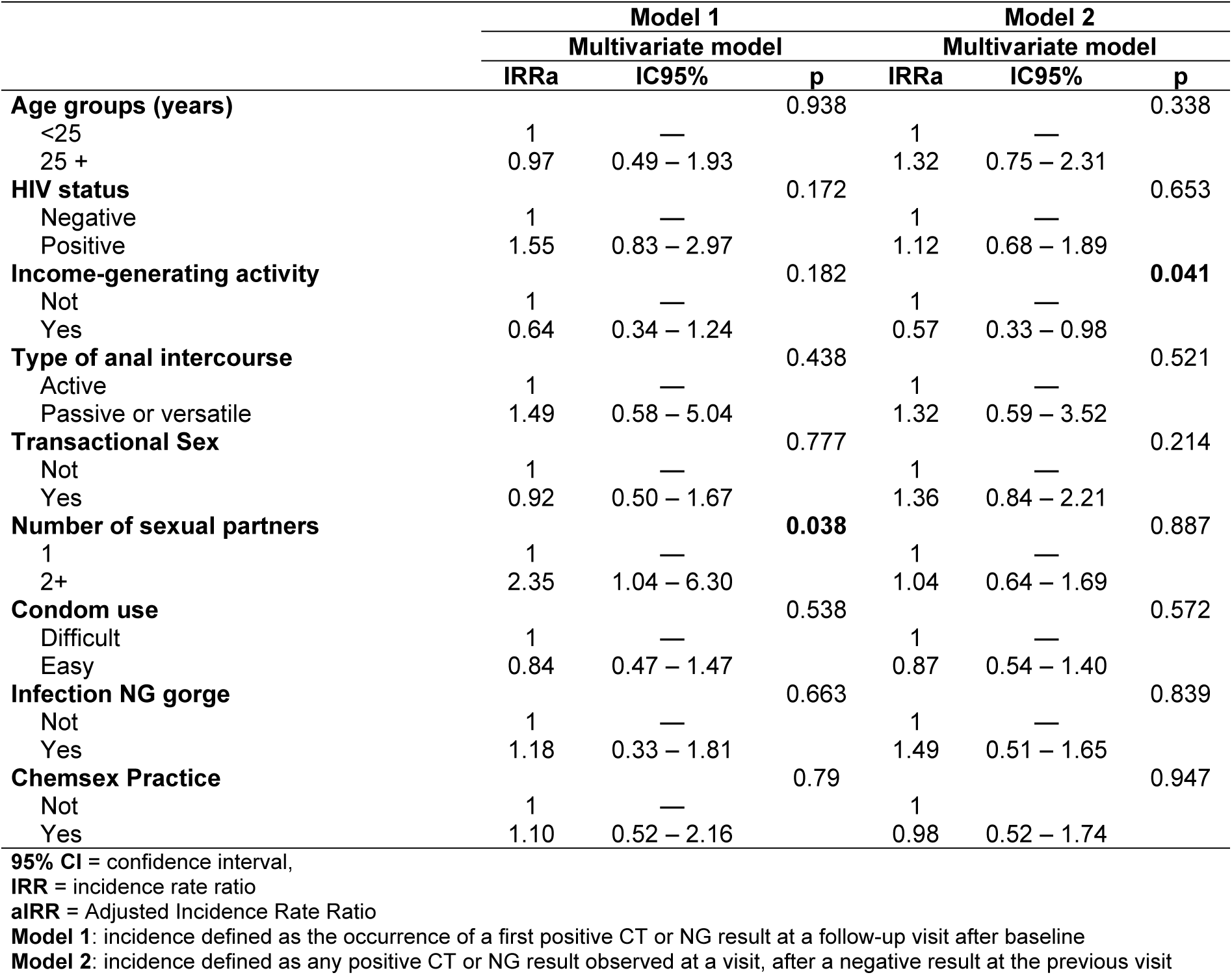
Factors associated with NG at the anal level, ANRS Cohort, DepIST H 12400, Togo, 2024.

|  | Model 1 |  |  | Model 2 |  |  |
| --- | --- | --- | --- | --- | --- | --- |
|  | Multivariate model |  |  | Multivariate model |  |  |
|  | IRRa | IC95% | p | IRRa | IC95% | p |
| **Age groups (years)** |  |  | 0.938 |  |  | 0.338 |
| <25 | 1 | — |  | 1 | — |  |
| 25 + | 0.97 | 0.49 – 1.93 |  | 1.32 | 0.75 – 2.31 |  |
| **HIV status** |  |  | 0.172 |  |  | 0.653 |
| Negative | 1 | — |  | 1 | — |  |
| Positive | 1.55 | 0.83 – 2.97 |  | 1.12 | 0.68 – 1.89 |  |
| **Income-generating activity** |  |  | 0.182 |  |  | **0.041** |
| Not | 1 | — |  | 1 | — |  |
| Yes | 0.64 | 0.34 – 1.24 |  | 0.57 | 0.33 – 0.98 |  |
| **Type of anal intercourse** |  |  | 0.438 |  |  | 0.521 |
| Active | 1 | — |  | 1 | — |  |
| Passive or versatile | 1.49 | 0.58 – 5.04 |  | 1.32 | 0.59 – 3.52 |  |
| **Transactional Sex** |  |  | 0.777 |  |  | 0.214 |
| Not | 1 | — |  | 1 | — |  |
| Yes | 0.92 | 0.50 – 1.67 |  | 1.36 | 0.84 – 2.21 |  |
| **Number of sexual partners** |  |  | **0.038** |  |  | 0.887 |
| 1 | 1 | — |  | 1 | — |  |
| 2+ | 2.35 | 1.04 – 6.30 |  | 1.04 | 0.64 – 1.69 |  |
| **Condom use** |  |  | 0.538 |  |  | 0.572 |
| Difficult | 1 | — |  | 1 | — |  |
| Easy | 0.84 | 0.47 – 1.47 |  | 0.87 | 0.54 – 1.40 |  |
| **Infection NG gorge** |  |  | 0.663 |  |  | 0.839 |
| Not | 1 | — |  | 1 | — |  |
| Yes | 1.18 | 0.33 – 1.81 |  | 1.49 | 0.51 – 1.65 |  |
| **Chemsex Practice** |  |  | 0.79 |  |  | 0.947 |
| Not | 1 | — |  | 1 | — |  |
| Yes | 1.10 | 0.52 – 2.16 |  | 0.98 | 0.52 – 1.74 |  |

95% CI = confidence interval,
IRR = incidence rate ratio
aIRR = Adjusted Incidence Rate Ratio
**Model 1:** incidence defined as the occurrence of a first positive CT or NG result at a follow-up visit after baseline**Model 2:** incidence defined as any positive CT or NG result observed at a visit, after a negative result at the previous visit

## Discussion

The objective of the present study was to estimate the incidence of bacterial STIs and to identify the associated factors among MSM in Lomé, Togo. Our results confirm that MSM remain a population at high risk of exposure to STIs, with incidence rates around 22 per 100 person-years for CT (21.8 cases per 100 person-years) and NG (22.0 cases per 100 person-years). Another concern is risky sexual behaviors, which are still prevalent in this community. In fact, in our study, only 46.0% of participants reported consistent condom use, 40.0% reported transactional sex, and 75.0% reported having multiple sexual partners. In addition, 20.0% of participants practice chemsex, which increases the risk of exposure.

In our study, the three anatomical sites explored (throat, urine, and anus) showed varying levels of positivity; the rectal area being the main site of carriage of these infections (CT (17.2 per 100 person-years) and NG (17.9 per 100 person-years)). Several studies have reported similar results, confirming the anal site as the main reservoir of STIs in MSM and its decisive role in transmission dynamics [10, 11]. In a study conducted in Kenya in 2020 on a sample of 104 MSM followed over a period of 9 months, the prevalence of anal STIs at baseline was particularly high, with one in five participants having an anal CT or NG infection with an incidence of around 30% [10]. The same observation was made in Tanzania, on a study of 300 MSM carried out on throat, anus and urine swabs. In this study, the rectal area was also the main site of carriage of CT and NG infections with more than 90% of the cases recorded [11]. One of the reasons given was the fact that the participants in the consultations only admit heterosexual intercourse to avoid stigmatization, which prevents any examination and diagnosis of rectal infection [11]. Also, rectal infections caused by CT and NG are most often asymptomatic, which further complicates diagnosis and management, especially in countries where a syndrome-based approach is still used in the management of STIs [12]. This explains why, in studies of these populations with systematic anal swabs, many positive results for STIs are recorded [11]. These findings are consistent with reports of high stigma of MSM by health professionals in sub-Saharan Africa [13]. In Togo, homosexuality is criminalized, which usually forces MSM to keep quiet about their sexual orientation so as not to be rejected [8]. They are victims of violence, particularly from those around them, but also of social discrimination in terms of employment and access to education or health care [14]. Despite the progress that has been made in the field of health, with the inclusion of MSM in the fight against HIV/AIDS and other STIs, this structural stigma reduces access to services among this population [15]. An online survey of MSM in 38 European countries found that in countries with more stigmatizing laws and policies, MSM were less likely to report same-sex attraction, sexual behaviors, and gender identity than in countries with less stigmatizing laws and policies. This non-disclosure of sexual behaviors has been shown to have a negative impact on the prevention, diagnosis and treatment of STIs [16]. All of these data thus underline the importance of decriminalization or the easing of repressive measures and the reduction of stigmatization, as essential elements of an effective fight against STIs in this population [17]. Community associations are strategic actors in the fight against STIs, especially in such a socio-political context marked by the stigmatization of MSM. Numerous studies show that community-based peer-based interventions play a key role in improving access to care, reducing risk, and adopting preventive behaviors among these groups [18]. Therefore, it would be relevant for decision-makers to promote the creation and strengthening of local associations involving MSM peers, to replicate these effective approaches in the country and thus fill the current gaps in sexual health.

In terms of associated factors, only MSM who reported multiple sexual partners had a significantly elevated risk of acquiring NG infection at the anal level. For HIV status, no significant association was found. This result can be explained by the fact that HIV-positive MSM in Togo benefit from enhanced monitoring, particularly in terms of sexual health.

In our study, a trend towards an increase in the incidence of CT at the anal level is observed in people with multiple partners (aIRR: 1.34; 95% CI: 0.66–2.96) but were not statistically significant. In Model 2, a similar dynamic appears: the incidence seems to increase in the presence of a pharyngeal infection (aIRR: 2.96; 95% CI: 0.73–8.78) or transactional reports (aIRR: 1.95; 95% CI: 0.89–4.30). However, the wide confidence intervals including 1 do not allow a significant association to be concluded. Regarding the factors associated with NG at the anal level, both models also suggest an increase in the incidence of pharyngeal infection. Although no statistically significant association was identified, these results suggest an increased risk of infection at one site if infection occurs at another. A Brazilian study conducted on 171 PrEP users showed that involvement of a urinary or oropharyngeal site generally precedes rectal involvement, given the sexual practices described within this community (oral or anal sex) [19]. In such a context, a major recommendation would be the implementation of triple screening or systematic multi-site screening (urinary, pharyngeal and anorectal) among MSM in Togo. Several studies have shown that this type of screening, even when clinical signs are present at only one site, would significantly increase the number of infections detected, especially asymptomatic STIs [20, 21]. This would allow effective management of these infections, in order to avoid developing a symptomatic STI, and break the chain of transmission [21]. However, this strategy represents a significant cost for health services, particularly in developing countries [22]. To address the cost issue, one option would be sample pooling. Indeed, the grouping of samples taken from the throat, urine and rectum between individuals has proven to be sensitive and specific for the detection of CT and NG and this could constitute an economical strategy with a minimal decrease in sensitivity. It increases the chances of detecting the pathogen while saving laboratory reagents and consumables [23, 24].

New strategies for preventing STIs are also being considered, including post-exposure prophylaxis with doxycycline. The latter has recently demonstrated its effectiveness in clinical trials conducted with MSM and transgender women in France and the United States, showing a two-thirds decrease in the incidence of chlamydia infections and early syphilis when doxycycline was taken within 72 hours of unprotected sex [25, 26]. These results argue in favor of its use in MSM who have recently contracted bacterial STI, but its effectiveness against gonorrhea could be reduced. Several vaccines targeting gonococcal disease are currently under investigation, and their use will be a major priority for sexual and reproductive health in low- and middle-income countries. One of the challenges in the coming years will be to re-evaluate the syndromic approach to STI management strategy or to integrate systematic treatment regardless of the presence of clinical symptoms [27, 28].

One of the main strengths of this study is that it provides incidence data and allows us to observe the dynamics of STIs among MSM while contributing to a better understanding of the associated factors in the Togolese context. Most of the previous work has focused on estimating STI prevalence without including follow-up. However, some limitations are worth noting. The first concerns the relatively small sample size. The basic assumptions for the calculation of the sample size were based on the estimation of the prevalence and incidence of anal lesions. This could explain the absence of a statistically significant association for certain variables usually reported in the literature. Another limitation is that the analysis strategy does not allow for changes in state during monitoring. Thus, infections that occurred and resolved between two follow-up visits may have compromised the accuracy of the estimates and influenced the results. However, these shortcomings do not affect the scope of the study, which will provide a solid basis for monitoring and response. In the future, the use of statistical models that consider state transitions would allow for a better understanding of infection dynamics and refine estimates.

## Conclusion

This study confirms that MSM remain highly exposed to STIs in Togo and highlights the need to strengthen prevention efforts within this population. Several strategies are currently being explored, including post-exposure prophylaxis with doxycycline despite the risk of resistance, as well as systematic multi-site screening with pooling of samples to optimize costs. Consideration of these approaches by policymakers will be essential to guide the development of future national recommendations.

## Data Availability

Data cannot be shared publicly because of their sensitivity (Data on HIV, bacterial sexually transmitted infections (STIs) and sexuality in a vulnerable population). Data are available from the Center for Training and Research in Public Health (Gerard KOGLO, Research Officer and Database Archiving Manager) for researchers who meet the criteria for access to confidential data.

## Abbreviations

aIRR: adjusted incidence rate ratio
CBRS: Bioethics Committee for Health Research
CI: Conficence interval
CT: Chlamydia trachomatis
IQR: Interquartile range
IRR: Incidence rate ratio
MSM: Men who have sex with men
NG: Neisseria gonorrhoeae
STI: Sexually transmitted infections
WHO: World Health Organization

## Declarations

## Ethics approval and consent to participate

The study involved human participants, and the protocol was approved by all participants, who provided written informed consent. The Bioethics Committee of the Ministry of Health in Togo (CBRS) approved the study protocol (N°039/2019/CBRS), and the study was registered on the ClinicalTrial.gov platform with the number NCT04910438.

## Consent for publication

Not applicable.

## Availability of data and material

The datasets used and/or analyzed during the current study are available from the corresponding author upon reasonable request.

## Authors’ contributions

A.J.S., V.M.F., D.D., D.K.E., and C.C. conceptualized the study. E.M. did the clinical visits and collected the samples. M.S. and A.C.D. performed and analyzed the laboratory analysis. O.I.W.A. and A.J.S drafted the analysis plan and wrote the first draft of the manuscript. O.I.W.A. performed the statistical analysis under the supervision of M.K.T and F.A.G.K. All authors contributed to data analysis through review and interpretation of the results. All authors read, revised and approved the final manuscript.

## Acknowledgements

We thank all the study participants and the staff of the Espoir Vie Togo NGO in Lomé, Togo. We thank Arabella Touati, Marie Gardette (CHU Bordeaux), and Mary Mainardis (St. Louis, Paris) for technical assistance.

## References

[1] World Health Organization (WHO). Sexually transmitted infections (STIs) : Fact sheets [Online]. 2025 [Cited by 2026 feb 08]. Available on : https://tinyurl.com/4fa34fry.

[2] Malekinejad M, Barker EK, Merai R, Lyles CM, Bernstein KT, Sipe TA, DeLuca JB, Ridpath AD, Gift TL, Tailor A, Kahn JG. Risk of HIV Acquisition Among Men Who Have Sex With Men Infected With Bacterial Sexually Transmitted Infections: A Systematic Review and Meta-Analysis. Sex Transm Dis. 2021 Oct 1;48(10):e138–e148. doi: 10.1097/OLQ.0000000000001403. PMID: 33783414; PMCID: PMC8485981.

[3] Blondeel K, Say L, Chou D, Toskin I, Khosla R, Scolaro E, Temmerman M. Evidence and knowledge gaps on the disease burden in sexual and gender minorities: a review of systematic reviews. Int J Equity Health. 2016 Jan 22;15:16. doi: 10.1186/s12939-016-0304-1. PMID: 26800682; PMCID: PMC4724086.

[4] Vallejo-Ortega MT, Gaitán Duarte H, Mello MB, Caffe S, Perez F. A systematic review of the prevalence of selected sexually transmitted infections in young people in Latin America. Rev Panam Salud Publica. 2022 Jun 21;46:e73. doi: 10.26633/RPSP.2022.73. PMID: 35747471; PMCID: PMC9211030.

[5] Atkins K, Wiginton JM, Carpino T, Sanchez TH, Murray SM, Baral SD. Transactional Sex, HIV, and Bacterial STIs Among U.S. Men Who have Sex with Men. Am J Prev Med. 2024 Nov;67(5):722–729. doi: 10.1016/j.amepre.2024.07.002. Epub 2024 Jul 11. PMID: 39002886.

[6] Baetselier ID, Crucitti T, Yaya I, et al P542 Prevalence of STIs among MSM initiating PrEP in west-africa (CohMSM-PrEP ANRS 12369 – expertise france) Sexually Transmitted Infections 2019;95:A245.

[7] Ferré VM, Sadio AJ, Gbeasor-Komlanvi FA, Bucau M, Salou M, Berçot B, Bébéar C, Abramowitz L, Zaidi M, Amenyah-Ehlan AP, Mensah E, Braille A, Couvelard A, Dagnra AC, Ghosn J, Descamps D, Charpentier C, Ekouevi DK. High prevalence of bacterial STI, anal HPV, cytological abnormalities and anal lesions among MSM in Togo, 2021: a baseline analysis of the ANRS I MIE 12,400/DepIST-H cohort. BMC Infect Dis. 2025 Sep 26;25(1):1156. doi: 10.1186/s12879-025-11338-y. PMID: 41013315; PMCID: PMC12465205.

[8] Togo. Loi n° 2015-010 du 27 octobre 2015 portant nouveau Code pénal de la République togolaise. Journal Officiel de la République Togolaise. 2015. Disponible sur : https://www.jo.gouv.tg/sites/default/files/publications/LOI_2015_010.pdf.

[9] Chan PA, Robinette A, Montgomery M, et al. Extragenital infections caused by Chlamydia trachomatis and Neisseria gonorrhoeae: a review of the infect dis. Obstet Gynecol. 2016;2016:5758387 PMID:27366021.

[10] Ngetsa CJ, Heymann MW, Thiong’o A, Wahome E, Mwambi J, Karani C, Menza NC, Mwashigadi G, Muturi MW, Graham SM, Mugo PM, Sanders EJ. Rectal gonorrhoea and chlamydia among men who have sex with men in coastal Kenya. Wellcome Open Res. 2020 Jun 4;4:79. doi: 10.12688/wellcomeopenres.15217.4. PMID: 32647750; PMCID: PMC7323594.

[11] Ross MW, Nyoni J, Ahaneku HO, Mbwambo J, McClelland RS, McCurdy SA. High HIV seroprevalence, rectal STIs and risky sexual behaviour in men who have sex with men in Dar es Salaam and Tanga, Tanzania. BMJ Open. 2014 Aug 28;4(8):e006175. doi: 10.1136/bmjopen-2014-006175. PMID: 25168042; PMCID: PMC4156794.

[12] Kent CK, Chaw JK, Wong W, Liska S, Gibson S, Hubbard G, Klausner JD. Prevalence of rectal, urethral, and pharyngeal chlamydia and gonorrhea detected in 2 clinical settings among men who have sex with men: San Francisco, California, 2003. Clin Infect Dis. 2005 Jul 1;41(1):67–74. doi: 10.1086/430704. Epub 2005 May 26.

[13] Lane T, Mogale T, Struthers H, McIntyre J, Kegeles SM. ‘They see you as a different thing’: the experiences of men who have sex with men with healthcare workers in South African township communities. Sex Transm Infect. 2008 Nov;84(6):430–3. doi: 10.1136/sti.2008.031567. PMID: 19028941; PMCID: PMC2780345.

[14] Afrique Arc-en-Ciel Togo, Synergía – Initiatives for Human Rights. Rapport alternatif soumis au Comité des droits de l’homme pour l’examen du Togo à la 132e session (28 juin–23 juillet 2021). Lomé (TGO): Afrique Arc-en-Ciel Togo; 2021. Disponible sur: https://ccprcentre.org/files/documents/INT_CCPR_CSS_TGO_45147_F.pdf.

[15] Cedoca. L’homosexualité [Internet]. Bruxelles: Commissariat général aux réfugiés et aux apatrides (CGRA); 2023 [cité 2026 janv 7]. Disponible sur: https://tinyurl.com/mtfkxnbf.

[16] Pachankis JE, Hatzenbuehler ML, Mirandola M, Weatherburn P, Berg RC, Marcus U, Schmidt AJ. The Geography of Sexual Orientation: Structural Stigma and Sexual Attraction, Behavior, and Identity Among Men Who Have Sex with Men Across 38 European Countries. Arch Sex Behav. 2017 Jul;46(5):1491–1502. doi: 10.1007/s10508-016-0819-y. Epub 2016 Sep 12. PMID: 27620320; PMCID: PMC5346459.

[17] Lyons CE, Twahirwa Rwema JO, Makofane K, Diouf D, Mfochive Njindam I, Ba I, Kouame A, Tamoufe U, Cham B, Aliu Djaló M, Obodou EP, Karita E, Simplice A, Nowak RG, Crowell TA, Matse S, Kouanda S, Enama JP, Kavanagh M, Millett GA, Beyrer C, Murray S, Baral S. Associations between punitive policies and legal barriers to consensual same-sex sexual acts and HIV among gay men and other men who have sex with men in sub-Saharan Africa: a multicountry, respondent-driven sampling survey. Lancet HIV. 2023 Mar;10(3):e186–e194. doi: 10.1016/S2352-3018(22)00336-8. Epub 2023 Jan 6. PMID: 36623537; PMCID: PMC10288909.

[18] Abubakari GM, Turner D, Ni Z, Conserve DF, Dada D, Otchere A, Amanfoh Y, Boakye F, Torpey K, Nelson LE. Community-Based Interventions as Opportunities to Increase HIV Self-Testing and Linkage to Care Among Men Who Have Sex With Men - Lessons From Ghana, West Africa. Front Public Health. 2021 Jun 11;9:660256. doi: 10.3389/fpubh.2021.660256. PMID: 34178919; PMCID: PMC8226123.

[19] Antonini M, Vettore MV, Øgård-Repål A, de Macêdo Rocha D, de Alencar Rocha KA, Elias HC, Barufaldi F, Santana RC, Gir E, Spire B, Reis RK. Patterns of Chlamydia trachomatis and Neisseria gonorrhoeae in different anatomical sites among Pre-Exposure Prophylaxis (PrEP) users in Brazil. BMC Infect Dis. 2024 Feb 26;24(1):260. doi: 10.1186/s12879-024-09144-z. PMID: 38408940; PMCID: PMC10895759.

[20] Zucker R, Gaisa M, Sigel K, Singer I, Adler A, Turner D, Ben Ami R, Nissan I, Chan C, Halperin T. Triple site sexually transmitted infection testing as a crucial component of surveillance for men who have sex with men: A prospective cohort study. Int J STD AIDS. 2022 Feb;33(2):114–122. doi: 10.1177/09564624211047477. Epub 2021 Oct 22. PMID: 34676780.

[21] VIH.org. Dépistage des IST asymptomatiques chez les HSH en 2024 : le point de vue des médecins en France [Internet]. Paris: VIH.org; 2024 août 28 [cité 2026 janv 7]. Disponible sur : https://tinyurl.com/5fba8mpe.

[22] Sultan B, White JA, Fish R, Carrick G, Brima N, Copas A, Robinson A, Gilson R, Mercey D, Benn P. The ‘3 in 1’ Study: Pooling Self-Taken Pharyngeal, Urethral, and Rectal Samples into a Single Sample for Analysis for Detection of Neisseria gonorrhoeae and Chlamydia trachomatis in Men Who Have Sex with Men. J Clin Microbiol. 2016 Mar;54(3):650–6. doi: 10.1128/JCM.02460-15. Epub 2015 Dec 30. PMID: 26719439; PMCID: PMC4767962.

[23] De Baetselier I, Osbak KK, Smet H, Kenyon CR, Crucitti T. Take three, test one: a cross-sectional study to evaluate the molecular detection of Chlamydia trachomatis and Neisseria gonorrhoeae in pooled pharyngeal, anorectal and urine samples versus single-site testing among men who have sex with men in Belgium. Acta Clin Belg. 2020;75(2):91–5. doi: 10.1080/17843286.2018.1545376.

[24] Verougstraete N, Verbeke V, De Cannière A-S, Simons C, Padalko E, Coorevits L. To pool or not to pool? Screening of Chlamydia trachomatis and Neisseria gonorrhoeae in female sex workers: pooled versus single-site testing. Sex Transm Infect. 2020;96:417–21. doi: 10.1136/sextrans-2019-054357.

[25] Palacios R, Gómez-Ayerbe C, Martín-Cortés S, González-Jiménez A, López-Jódar M, Pérez-Hernández IA, Santos J. Post-exposure prophylaxis with doxycycline (DoxyPEP) to prevent STIs in a real-world setting: the PRIDOX study. J Antimicrob Chemother. 2025 Dec 2;80(12):3306–3310. doi: 10.1093/jac/dkaf366. PMID: 41189494; PMCID: PMC12670152.

[26] Molina JM, Bercot B, Assoumou L, Rubenstein E, Algarte-Genin M, Pialoux G, Katlama C, Surgers L, Bébéar C, Dupin N, Ouattara M, Slama L, Pavie J, Duvivier C, Loze B, Goldwirt L, Gibowski S, Ollivier M, Ghosn J, Costagliola D; ANRS 174 DOXYVAC Study Group. Doxycycline prophylaxis and meningococcal group B vaccine to prevent bacterial sexually transmitted infections in France (ANRS 174 DOXYVAC): a multicentre, open-label, randomised trial with a 2 × 2 factorial design. Lancet Infect Dis. 2024 Oct;24(10):1093–1104. doi: 10.1016/S1473-3099(24)00236-6. Epub 2024 May 23. PMID: 38797183.

[27] Donenberg GR, Kendall AD, Emerson E, Fletcher FE, Bray BC, McCabe K. IMARA: A mother-daughter group randomized controlled trial to reduce sexually transmitted infections in Black/African-American adolescents. PLoS One. 2020 Nov 2;15(11):e0239650. doi: 10.1371/journal.pone.0239650. PMID: 33137103; PMCID: PMC7605636.

[28] Gilbert L, Goddard-Eckrich D, Chang M, Hunt T, Wu E, Johnson K, Richards S, Goodwin S, Tibbetts R, Metsch LR, El-Bassel N. Effectiveness of a Culturally Tailored HIV and Sexually Transmitted Infection Prevention Intervention for Black Women in Community Supervision Programs: A Randomized Clinical Trial. JAMA Netw Open. 2021 Apr 1;4(4):e215226. doi: 10.1001/jamanetworkopen.2021.5226. PMID: 33835175; PMCID: PMC8035652.

